# Mortality trends and lengths of stay among hospitalized COVID-19 patients in Ontario and Québec (Canada): a population-based cohort study of the first three epidemic waves

**DOI:** 10.1101/2021.12.07.21267416

**Authors:** Yiqing Xia, Huiting Ma, David L Buckeridge, Marc Brisson, Beate Sander, Adrienne Chan, Aman Verma, Iris Ganser, Nadine Kronfli, Sharmistha Mishra, Mathieu Maheu-Giroux

**Affiliations:** Department of Epidemiology and Biostatistics, School of Population and Global Health, McGill University, Montréal, QC, Canada; MAP Centre for Urban Health Solutions, St. Michael’s Hospital, Unity Health Toronto, Toronto, ON, Canada; Département de médecine sociale et préventive, Faculté de médecine, Université Laval, Québec, QC, Canada; Toronto Health Economics and Technology Assessment (THETA) collaborative, University Health Network; Institute of Health Policy, Management and Evaluation (IHPME), Dalla Lana School of Public Health, University of Toronto; Public Health Ontario, Toronto, ON, Canada; ICES, Toronto, ON, Canada; Division of Infectious Diseases, Department of Medicine, University of Toronto, Toronto, ON, Canada; Centre for Outcomes Research and Evaluation (CORE), Research Institute of the McGill University Health Centre, Montréal, QC, Canada; Department of Medicine, Division of Infectious Diseases and Chronic Viral Illness Service, McGill University Health Centre, Montréal, QC, Canada; Institute of Medical Sciences, University of Toronto

## Abstract

**Background:** Epidemic waves of COVID-19 strained hospital resources. We describe temporal trends in mortality risk and length of stay in intensive cares units (ICUs) among COVID-19 patients hospitalized through the first three epidemic waves in Canada.

**Methods:** We used population-based provincial hospitalization data from Ontario and Québec to examine mortality risk and lengths of ICU stay. For each province, adjusted estimates were obtained using marginal standardization of logistic regression models, adjusting for patient-level characteristics and hospital-level determinants.

**Results:** Using all hospitalizations from Ontario (N=26,541) and Québec (N=23,857), we found that unadjusted in-hospital mortality risks peaked at 31% in the first wave and was lowest at the end of the third wave at 6-7%. This general trend remained after controlling for confounders. The odds of in-hospital mortality in the highest hospital occupancy quintile was 1.2 (95%CI: 1.0-1.4; Ontario) and 1.6 (95%CI: 1.3-1.9; Québec) times that of the lowest quintile. Variants of concerns were associated with an increased in-hospital mortality. Length of ICU stay decreased over time from a mean of 16 days (SD=18) to 15 days (SD=15) in the third wave but were consistently higher in Ontario than Québec by 3-6 days.

**Conclusion:** In-hospital mortality risks and lengths of ICU stay declined over time in both provinces, despite changing patient demographics, suggesting that new therapeutics and treatment, as well as improved clinical protocols, could have contributed to this reduction. Continuous population-based monitoring of patient outcomes in an evolving epidemic is necessary for health system preparedness and response.

## Introduction

The COVID-19 pandemic has put immense pressure on health care systems. Canada’s most populous provinces, Ontario and Québec, bore the brunt of the pandemic^1^. These two provinces accounted for 70% of the country’s total number of COVID-19 hospitalizations during the first three epidemic waves^2,3^. The prolonged surges in hospital admissions led to rapid increases in hospital occupancy, especially in intensive care units (ICUs), with associated cancellations of non-urgent care^4-7^.

In-hospital mortality provides a proxy measure of the severity of a pandemic and the quality and effectiveness of hospital care ^8^. Worldwide, in-hospital mortality was highest in the first months of the pandemic, but progressively declined afterward^9,10^. Reasons for this decline include changes in who became infected (patient-level characteristics such as age and comorbidities)^11,12^, incremental improvements in clinical practice and treatment regimens ^13^, and refinement of critical care capacity^14,15^. However, the evolution of in-hospital mortality across the three epidemic waves has yet to be systematically examined in Canada, and it remains unclear to what extent these different factors might explain changes in the risk of COVID-19 in-hospital mortality in Canada.

During the course of the pandemic, projections of future demands for hospital beds have helped decision-makers to appropriately manage and allocate limited healthcare resources^16-18^. Predicting those demands requires an estimate of both the number of incoming patients needing different levels of hospital care and how long each person will stay in hospital^19^. Understanding the drivers of in hospital COVID-19 mortality is important to improve the accuracy of those projections. In addition, the disproportionate needs for care in ICUs warrants a thorough investigation of temporal changes in length of ICU stays^20,21^. The latter is a key metric that provides information on likely healthcare burden^19^.

In North America, most studies of in-hospital COVID-19 mortality were informed by the experiences of a single city^22,23^ or hospital^1,24^ and the generalizability of these findings remains unclear. To address the above knowledge gaps, we conducted a population-based cohort study using data from the two largest Canadian provinces, Ontario and Québec, where over 60% (23.5 million) of the Canadian population reside. Specifically, our aims are to 1) describe temporal trends in in-hospital COVID-19 mortality risk, 2) understand drivers of changes in mortality risk, and 3) estimate changes in length of ICU stays over time.

## Methods

### Study design and setting

We conducted a retrospective population-based cohort study using provincial COVID-19 hospitalization databases from Ontario and Québec. Both provinces have a universal health care system and these databases capture all hospitalizations, with no loss to follow-up. Healthcare is under provincial jurisdiction, however, and the magnitude of epidemic waves and clinical protocols for COVID-19 patients may differ across provinces.

### Cohort eligibility criteria

Cohort entry occurs when individuals are admitted to an Ontario or Québec hospital with a COVID-19 diagnosis or the date of diagnosis that occurs a week or more after admission (i.e., presumed nosocomial infections considering the incubation period of COVID-19^25^). We included all hospitalizations with a lab-confirmed COVID-19 diagnosis admitted between March 1^st^ 2020 and May 31^st^ 2021 in Ontario and Québec. All observations were censored at discharge, death, or on August 15^th^ 2021, whichever occurred first. Among the very few individuals that experienced re-infection, only hospitalizations related to the first lab-confirmed episode were included in Québec as reinfections are milder than primary infections^26^. It was not possible to differentiate between initial and re-infection in Ontario database. Patients admitted or tested after discharge were excluded. Participants with missing date of ICU discharge were excluded from the analyses of lengths of stay (**Figure 1**).

**Figure 1.**
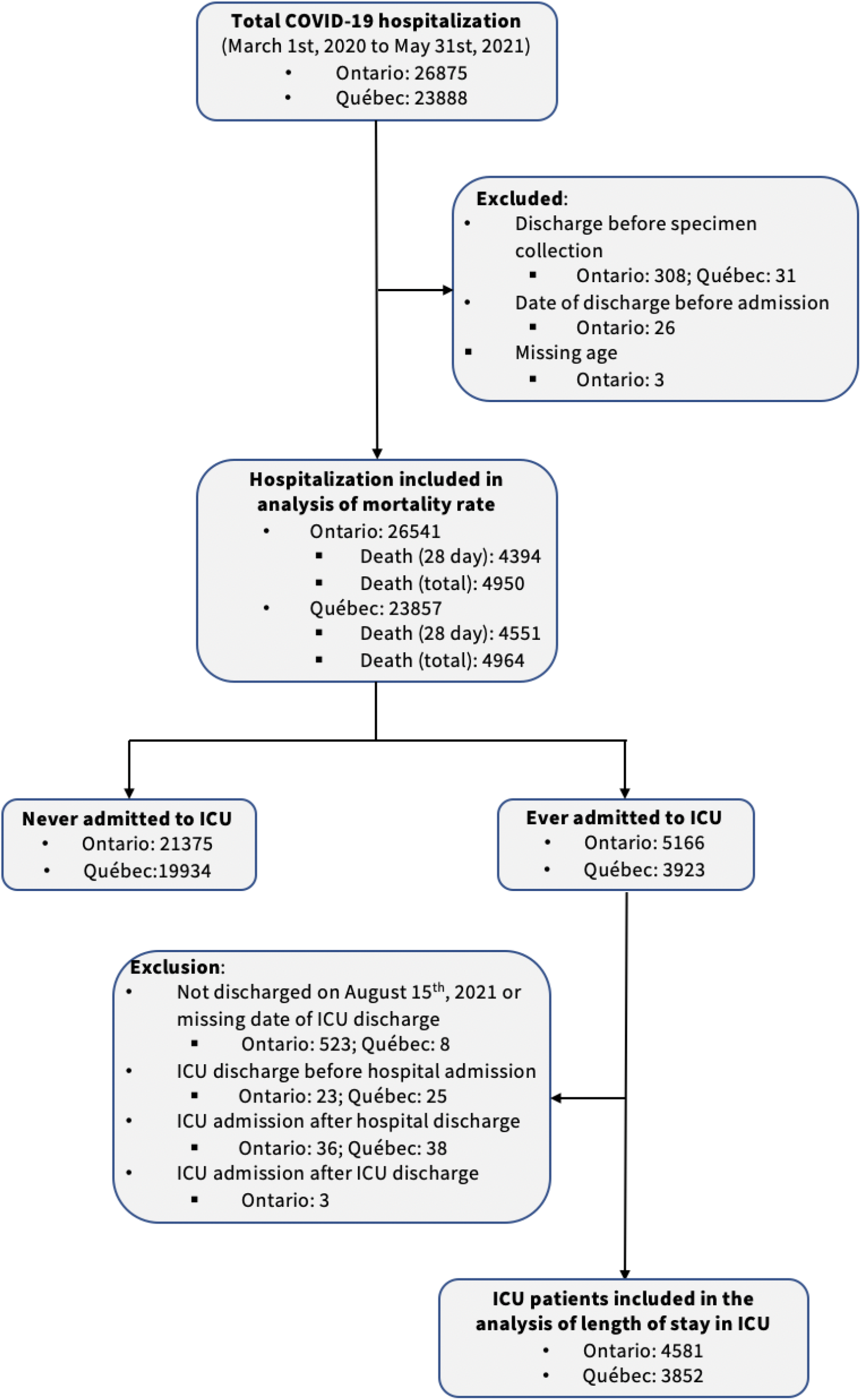
Flowchart of patients hospitalized with a lab-confirmed SARS-CoV-2 diagnosis included in the different analyses, by province (March 1^st^ 2020 to May 31^st^ 2021).

### Data sources

The Ontario datasets were made available by the Ontario Ministry of Health. Hospitalization data for Ontario was obtained from the *Ontario’s Case and Contact Management* (CCM+), a provincial surveillance database for reporting *Diseases of Public Health Significance*. The approaches dealing with missing observations in CCM+ reports were summarized in *Supplementary Text1*. Information on daily hospital capacity at the level of the province’s 34 public health units came from *Bed census summary dataset*.

In Québec, data were obtained from the *Maintenance et exploitation des données pour l’étude de la clientèle hospitalière* database (MED-ÉCHO live). Daily hospital capacity data were abstracted from the *Relevé quotidien du centre hospitalier*, which contains the number of regular/ICU beds available and occupied at each hospital. All Québec datasets came from Québec *Health Insurance databases* (*Régie de l’assurance maladie du Québec, RAMQ*) and databases from the Québec Ministry of Health and Social Services (*Ministère de la Santé et des Services sociaux, MSSS*). The access to these databases is made possible through a tripartite agreement between the MSSS, the RAMQ, and the *Institut national d’excellence en santé et en services sociaux*.

### Outcome and variables

Two primary outcomes were studied. The first was all-cause in-hospital mortality, defined as a death occurring within 28 days of admission, in line with other studies^27-30^. Patients discharged or died after 28 days were coded as *alive* at 28 days. The other outcome of interest was the length of stay in ICU, defined as the time from ICU admission to ICU discharge or death (inclusive of the latter).

We categorized patients based on their admission date and corresponding epidemic wave: *Wave 1* (before August 1^st^, 2020), *Wave 2* (August 1^st^, 2020 to March 20^th^, 2021), and *Wave 3* (after March 20^th^, 2021). Hospitalizations with a first positive specimen collected 7 days or more after the date of admission, or whose living environment is the hospital, were classified as *hospital-acquired infection*. In such cases, the date of hospital admission for COVID-19 was replaced with the date of the first positive specimen to better reflect the time of infection. Patients who were admitted to ICU the same day as their hospital admission were defined as *direct ICU admission*. Patients positive for a variant of concern (VOC), through screening of specific mutations (N501Y and E484K in Ontario; N501Y, del69/70, and E484K in Québec), were regarded as *VOC positive* (mostly B.1.1.7, with some B.1.351 and P.1). Only those whose specimen collection date were 14 days after second dose were treated as *vaccinated* (only available in Québec by linking the provincial vaccine registry and MED-ÉCHO databases). *Overall hospital COVID-19 occupancy rate* was calculated for each day using the number of COVID-19 patients currently hospitalized as numerator and bed capacity (regular+ICU) as the denominator. This occupancy variable was categorized into quintiles from the lowest to highest independently for each province. In Québec, the occupancy rate was determined at the level of 88 hospitals, while in Ontario the variable was estimated at the level of 34 public health units due to data limitations. The *ICU occupancy rate* was calculated based on the same algorithm but using ICU bed capacity as the denominator. The gender of hospitalized patients (as proxy of biological sex) was not available in Québec.

### Statistical analyses

Unadjusted weekly mortality rates, stratified by age, wave, and quintiles of occupancy were calculated as the proportion of patients admitted with COVID-19 that deceased within 28 days for each time period and group. Uncertainty was quantified using 95% Clopper-Pearson confidence intervals (CI). Generalized linear models were used to fit smoothed curves of the weekly mortality rate (with regression cubic spline for the week of admission) and the mortality rates by age and wave. All analyses were performed for each province separately.

Adjusted estimates of mortality risk were obtained using logistic regression models that included cubic splines for calendar time (three knots). In addition, the models adjusted for patient-level characteristics and hospital-level determinants. Patient-level variables included those associated with severe outcome: age (cubic spline with knots at 50, 70, and 80 years; chosen using Akaike Information Criterion), gender (in Ontario), whether the patient was a resident of long-term care homes (LTCH), hospital-acquired infections status, direct admission to ICU, VOC status, and vaccination status (in Québec)^1,30-33^. Hospital-level determinants comprise the COVID-19 hospital occupancy rate (quintile ranking) at time of admission^34^ and facility-level fixed effects to control for time-invariant measured or unmeasured confounders. Because of lack of data disaggregation in Ontario, both occupancy and region fixed-effects were included at the public health unit level. Marginal standardization was used to obtain adjusted mortality risks over time, standardizing over all hospitalized patients. The 95%CI for overall adjusted mortality rates were generated using 1,000 bootstrap replicates.

Finally, we examined change in ICU length of stay across patient-level and hospital/region characteristics. Specifically, we calculated mean and standard deviation (SD) and used Kaplan-Meier stratified by age groups (0-49, 50-59, 60-69, 70-79, 80 years and older), by waves, and by ICU occupancy quintiles. The statistical significance of differences between survival curves was assessed using log-rank tests.

### Ethics approval

Ethics approvals were obtained from the *Health Sciences Research Ethics Board* of University of Toronto (no. 39253) in Ontario, and the *Institutional Review Board* of Faculty of Medicine and Health Sciences of McGill University in Québec (A06-M52-20B).

## Results

There were 26,541 (Ontario) and 23,857 (Québec) COVID-19 hospitalizations during the study period. Among them, 4,950 (Ontario) and 4,964 (Québec) deceased. Most of the deaths occurred within 28 days of admissions: 4,394 (89%) in Ontario and 4,551 (92%) in Québec. Nearly a fifth of patients were admitted to ICU during their hospital stay: 5,166 (20%) in Ontario; 3,923 (16%) in Québec. Overall hospital COVID-19 occupancy ranged from 0% to 47 % (Q1: 4%; Q2: 7%; Q3: 11%) in Ontario and from 0% to 51% (Q1: 5%; Q2: 10%; Q3: 16%) in Québec. Occupancy rate in ICU varied between 0% to 83% in Ontario (Q1: 3%; Q2: 7%; Q3: 11%) and between 0% to 123% (Q1: 13%; Q2: 22%; Q3: 33%) in Québec.

Hospitalization profiles varied over time: patients admitted during the third wave were younger than those admitted during the first two waves (**Table 1**). Patients with presumed hospital-acquired infection, those admitted directly to ICU, those who were not fully vaccinated, and those infected with a VOC were more likely to die in hospital. A decrease in the proportion of hospitalizations transferred from LTCH occurred after the first wave. Patients transferred from LTCH experienced higher mortality rates throughout the whole study period in Ontario. In Québec, however, LTCH patients were less likely to die in hospitals during the third wave, reflecting partly changes in directives between waves related to these transfers.

**Table 1.**
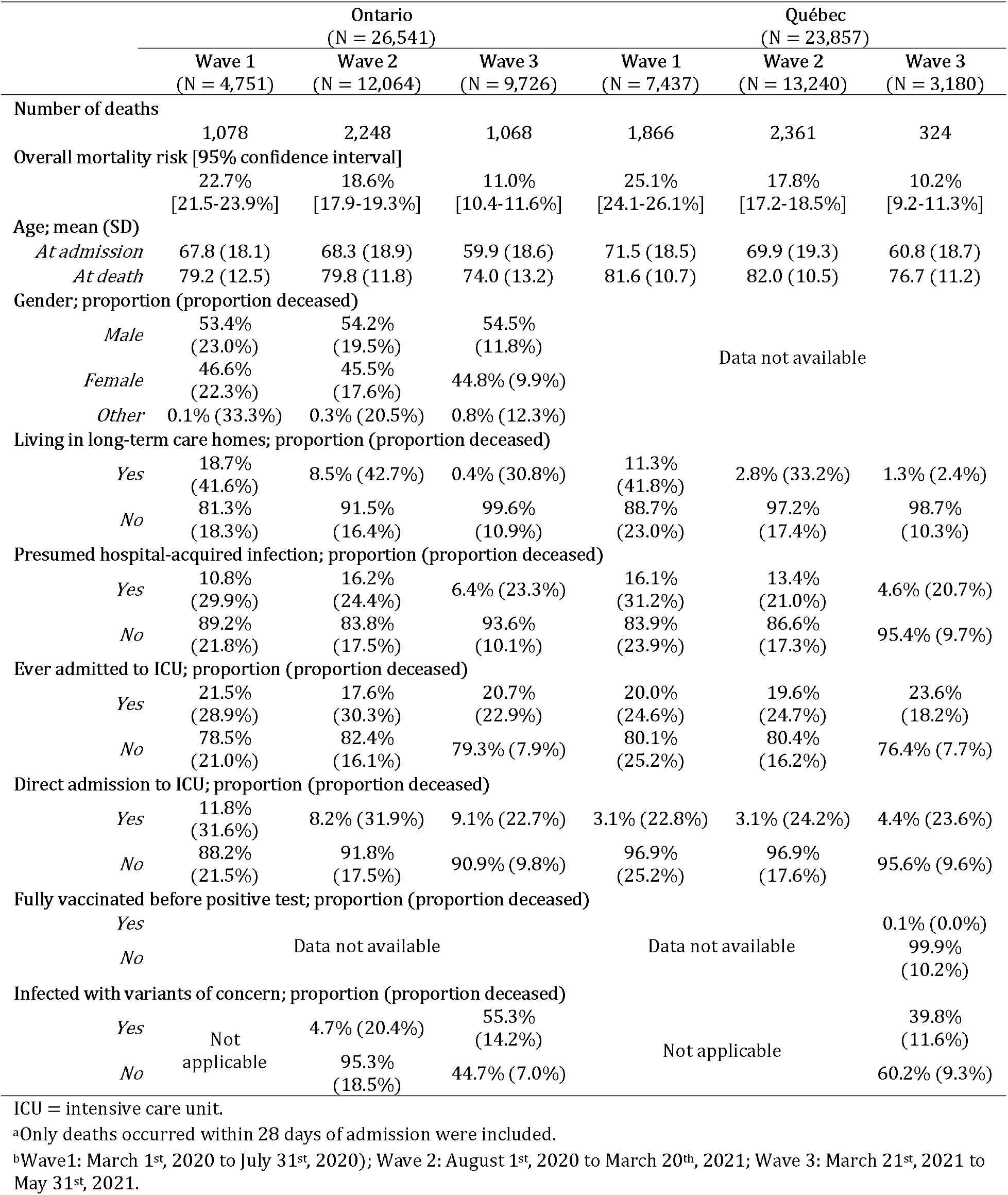
Characteristics of patients hospitalized with a laboratory-confirmed SARS-CoV-2 diagnosis and proportion deceased in Ontario and Québec (March 2020 to May 2021).

### Time trends in crude mortality risk among hospitalized COVID-19 patients

The time trends in crude mortality risk were similar between provinces (**Figure 2**). In the first two months of the epidemic, the probability of in-hospital death peaked at 31% (95%CI: 27-35%) in Ontario and 31% (95%CI: 28-34%) in Québec; followed by a gradual decrease in mortality that lasted until the beginning of the second wave. Thereafter, the risk of in-hospital death gradually increased, but plateaued at lower levels than in the first wave at 23% (95% CI: 20-27%) in Ontario and 23% (95% CI: 19-27%) in Québec. Starting from the middle of the second wave, a small decline occurred until the end of the third wave in both provinces. Overall, the unadjusted mortality rate followed the number of new hospitalizations, except for the third wave when mass vaccination was taking place.

**Figure 2.**
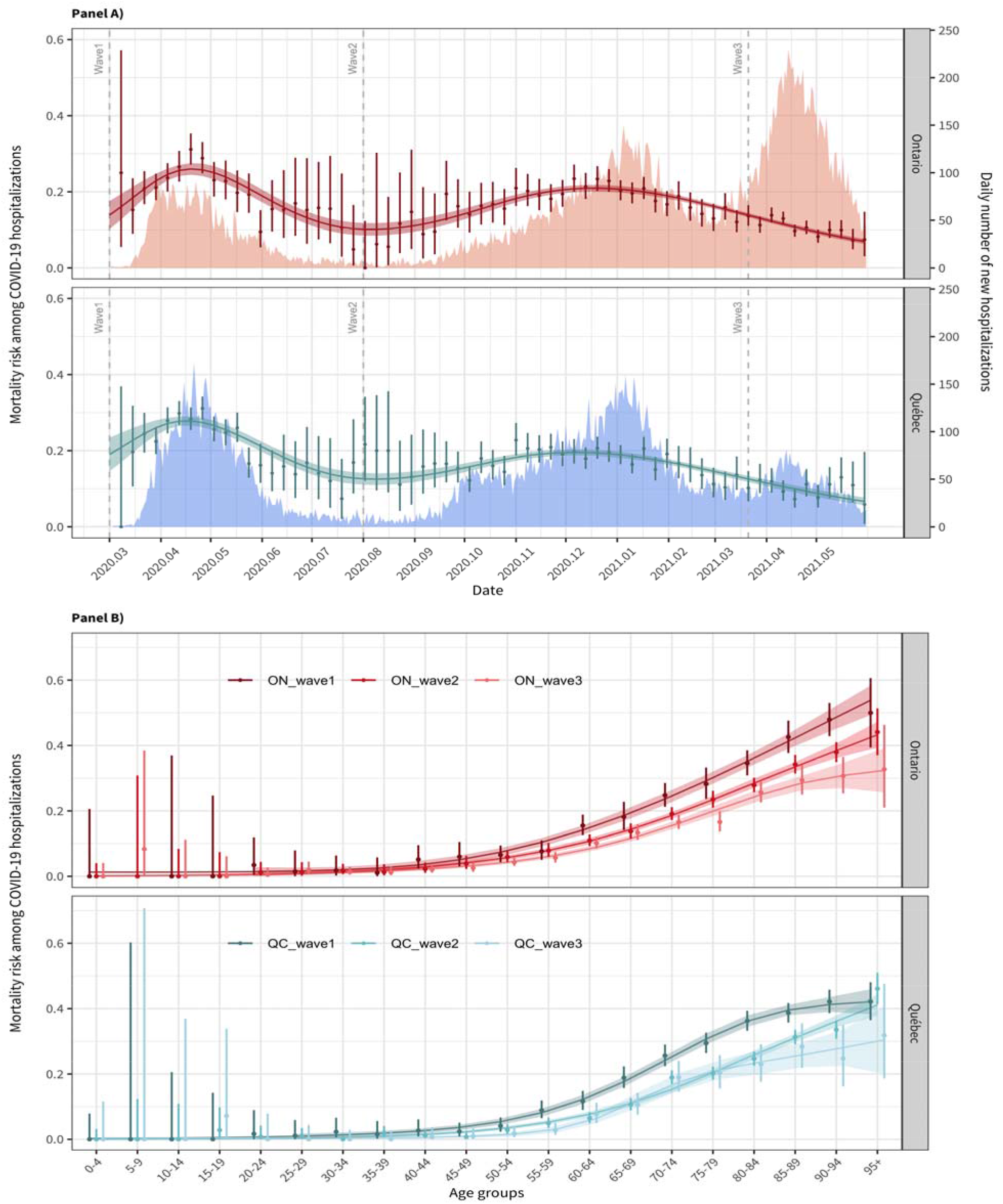
**Panel A:** Unadjusted weekly mortality risk among patients hospitalized with COVID-19 in Ontario and Québec. Point estimates are presented with 95% Clopper-Pearson confidence intervals. Mortality for the first week of March 2020 is not presented as only 5 and 1 patients were admitted in Ontario and Québec, respectively. Fitted mortality risk over time using binomial logistic regression models with cubic splines for week of admission are shown (curves) with associated confidence intervals (shaded areas around the curve). Daily numbers of new patients hospitalized with a COVID-19 diagnosis were presented as the shaded background. **Panel B**: Unadjusted mortality risk among patients hospitalized with COVID-19 by 5-year age groups, stratified by epidemic waves, with 95% Clopper-Pearson confidence intervals in Ontario and Québec. For each age group, mortality risks during Wave 1 (before August 1^st^, 2020), Wave 2 (August 1^st^, 2020 to March 20^th^, 2021), and Wave 3 (March 21^st^, 2021 to May 31^st^, 2021) are shown separately in that order from left to right. There was no hospitalization aged 5-9 years in Ontario during Wave 1.

There was a strong gradient in mortality risk with age in both provinces, and generally, the absolute mortality risk decreased over time for all age groups (**Figure 2**). However, there may have been less of a difference among the 60-84 group between the second and third wave in Québec. Hospital-level characteristics such as occupancy were also associated with mortality risk in crude analyses in Québec: there was a monotonic increase in crude mortality risk with increasing quintiles of facility-level occupancy (**Figure S1**). The trend in Ontario, where occupancy was measured at the level of the PHU, was stable through occupancy quintiles.

### Adjusted mortality risk over time

After adjusting for age, living environment, hospital-acquired infection status, direct ICU admission, infection with a VOC, vaccination status (in Québec only), and time-varying quintiles of hospital occupancy, the estimated temporal trend in mortality risk was similar to the unadjusted ones in both provinces (**Figure 3**). Despite this, Québec exhibited a more pronounced decrease in the estimated mortality risk at the beginning of the epidemic: from 37.1% (95% CI: 27.7-45.8%) to 15.2% (95% CI: 13.2-17.4%). In Ontario, the estimated decline for the same time period was from 24.7% (95% CI: 18.7-31.6%) to 13.5% (95% CI: 11.3-16.0%). Adjusted highest mortality risks during the second wave were comparable in Ontario (18.9%; 95% CI: 18.0-19.8%) and in Québec (18.2%; 95% CI: 17.3-19.0%) but the decline in the third wave was more pronounced in Ontario.

**Figure 3.**
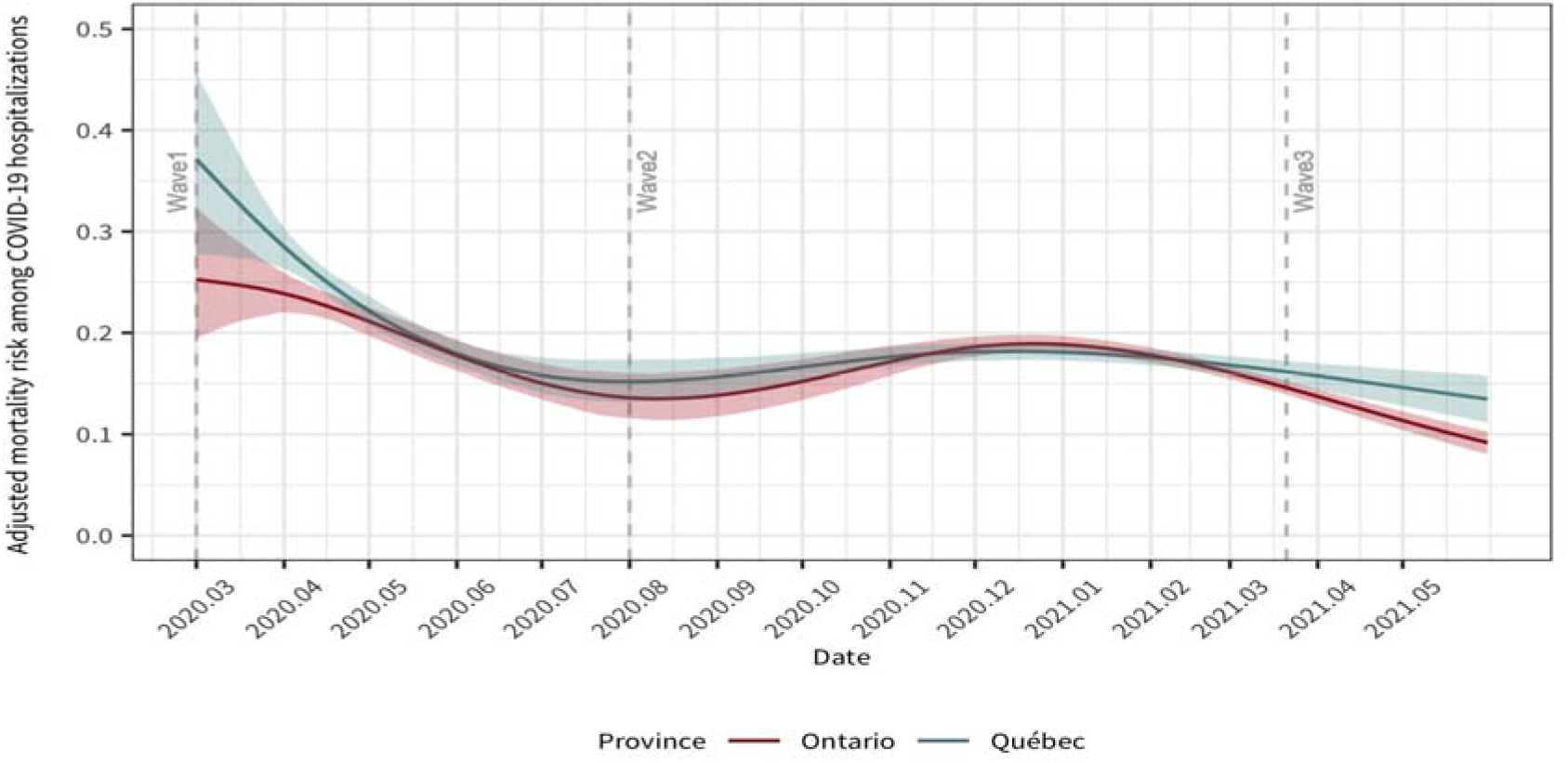
Adjusted mortality risk among patient hospitalized with COVID-19 and 95% bootstrapped confidence intervals in Ontario (in red) and Québec (in blue) since March 1^st^, 2020. The models were adjusted for quintile of occupancy at time of admission, age (cubic spline with 3 knots at 50, 70, and 80 years), gender (in Ontario), whether the patient was from long-term care home, had an hospital-acquired infection, direct admission to the intensive care unit, infection with a variant of concern, full vaccination status (in Québec), and either facility-level fixed effects (in Québec) or public health unit-level fixed effects (in Ontario). The absolute adjusted mortality risks were obtained by marginalizing over each province patient’s characteristics, which respective distributions differ slightly.

In adjusted analyses, mortality risk was higher at the second and highest occupancy quintile in Ontario, while it increased with the occupancy rate in Québec. The adjusted odds of in-hospital mortality in the highest occupancy quintile were 1.2 (95%CI: 1.0-1.4) and 1.6 (95%CI: 1.3-1.9) times that of the lowest one in Ontario and Québec respectively. In addition, the odds among patients that were male (adjusted OR: 1.4; 95%CI: 1.3-1.5, as compared to females), resident of LTCH (aOR=2.4 in Ontario, 95%CI: 2.1-2.7; aOR=1.7 in Québec, 95%CI: 1.5-2.0), with presumed hospital-acquired infections (aOR=1.5 in Ontario, 95%CI: 1.4-1.7; aOR=1.0 in Québec, 95%CI: 1.0-1.2), directly admitted to the ICU (aOR=3.7 in Ontario, 95%CI; 3.3-4.1; aOR=2.5 in Québec, 95%CI: 2.1-3.1), or were infected with a VOC (aOR=2.0 in Ontario, 95%CI: 1.7-2.3; aOR=1.3 in Québec, 95%CI: 1.0-1.7), were higher in both provinces. In Québec, none of the fully vaccinated COVID-19 hospitalized patients died. Odds ratio for these covariates are summarized in **Table S1**.

### Lengths of stay in intensive care units

Over the whole study period, the average length of stay in ICU was longer in Ontario (17.2 days) as compared to Québec (12.9 days; p-value<0.01). This trend was observed for all age groups (all p-values<0.01; **Table S2**). Comparing the three waves, lengths of stay in ICU decreased steadily over time in Ontario from 19.4 days (first wave), 17.6 days (second wave), to 15.6 days (third wave; all pairwise p-values<0.01; **Figure 4**). In Québec, average length of stays in ICU were 13.5 days during the first wave and then stabilized at 12.6 days in the following two waves. Age was associated with length of stays in both province: hospitalized individuals aged 0 to 49 years and those aged 80 years and older spend less time in ICU than others age groups (all pairwise p-values<0.01). The age-specific pattern was generally consistent across epidemic waves (**Figure S2**). In addition, there was a trend of shorter length of stays in ICU among hospitalized patients younger than 70 years of age with each epidemic wave in Ontario (p-value<0.01; **Figure S3**). No conclusive pattern was observed for the length of stays by ICU occupancy rate (**Figure S4, S5**). Patients who died and those who never used ventilator spent less time in ICU (p-value<0.01, except for the third wave; **Figure S6-S7**).

**Figure 4.**
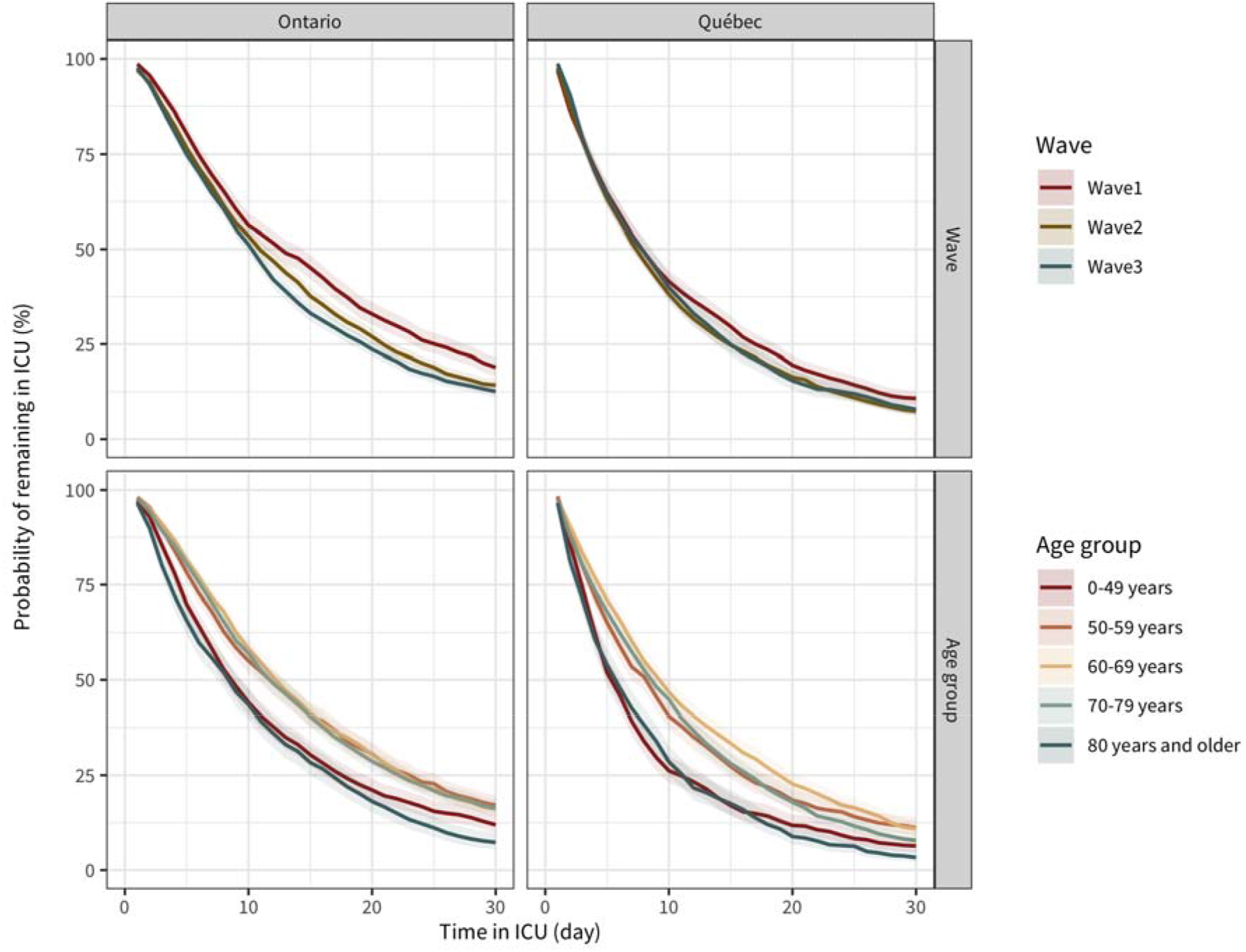
Kaplan-Meier curves for lengths of stay in intensive care units (ICU) among patients hospitalized with COVID-19, stratified by age group and by wave, in Ontario (top row) and Québec (bottom row). Waves are defined as followed: first wave1 (before June 30^th^, 2020), second wave (August 23^rd^, 2020 to March 20^th^, 2021), and third wave (March 21^st^, 2021 to May 31^st^, 2021).

## Discussion

Using population-based provincial surveillance databases containing records of all hospitalized COVID-19 patients in the two largest Canadian provinces, this study found important variations in mortality risk. Part of the observed decline over the three epidemic waves could be explained by changes in patient characteristics. Specifically, we found that the demographic profile of those acquiring infection (e.g., age, LTCH residents), hospital-acquired infections, VOC, and higher occupancy rates were associated with higher mortality risk. During periods of highest patient load (higher occupancy), the adjusted in-hospital mortality increased in both provinces. The length of ICU stay was consistently longer in Ontario compared to Québec. Patients aged 0-49 years and those 80 years and older were discharged from ICU more rapidly.

The observed substantial decrease in mortality risk during the first wave in both provinces is consistent with results from studies in the United Kingdom and the United States that adopted the same definition of in-hospital death^29,35,36^. Furthermore, given the discrepant epidemiological curves between Ontario and Québec, the similarity in the adjusted temporal trends in mortality risk also provides evidence that factors beyond patient profiles could have played a role. Reasons behind the persistent reduction in mortality could include adoption of new therapeutics and treatments. For example, dexamethasone and anti-IL-6 receptor monoclonal antibodies, which have been shown to reduce mortality among severely ill patients in the RECOVERY trial^28^ became part of treatment guidelines in early summer of 2020. Other potential factors include the cumulative experiences of hospital teams and the availability of updated evidence-based COVID-19 protocols^35,37,38^. The availability of COVID-19 vaccines in the third wave may also contributed to the continuous decreasing mortality rate with increasing coverage of first dose vaccine during that period^39^.

Overall, our analyses suggest that part of these in-hospital mortality risk reductions could be sustained if hospital capacity is maintained and hospital-acquired infections are prevented. These findings are aligned with those from studies conducted worldwide ^14,25,40-42^. Limited critical care resources and rapidly increasing staff-to-patient ratio could have influenced patient outcomes during periods of high transmission^29,43^. Additionally, nosocomial infections could exacerbate mortality risk because the already admitted patient population has vulnerable health conditions and higher rates of comorbidities^42,44^.

Concomitant with reductions in mortality risks, decreases in the length of ICU stays have been observed in multiple settings during the first wave^45,46^. Our results suggest a continuous decline in ICU stay throughout the study period. Despite the similar temporal patterns between provinces, we observed that the length of ICU stay in Ontario was consistently longer than it in Québec, and the proportion of patients admitted to ICU was higher in Ontario as well. Inter-provincial differences in clinical practices, such as criteria for ICU admission and discharge, could explain part of these differences. Other reasons include the changing demographic profiles of COVID-19 admissions. For example, patients aged 0 to 49 years and those 80 years and older spent less time in ICU than the others. Potentially because younger patients (≤50 years) improve more rapidly^47^ and those in the oldest age group experience higher mortality in ICU^48,49^. These findings are consistent with the observed shorter ICU stay among those who died and those who never used ventilator.

Tracking the evolution of patient outcomes can help improve hospital services, supply chain management, human resources planning, and prioritize future research^51^. In addition, the average length of ICU stay is a critical metric required to project census ICU bed, which has been a limiting factors of healthcare systems in several settings^52^. Despite differences in the proportion of patients admitted to ICU and their length of stay, the in-hospital mortality risks were relatively consistent between Ontario and Québec. Improving our understanding of ICU demand may contribute to optimizing patient outcomes and help planning for sufficient hospital capacity to adapt to potential increases in patient flow^14,53^.

Our study should be interpreted considering certain limitations. First, we were unable to control for sex^54^, ethnicity^55,56^, or comorbidities^57^—factors that have been associated with COVID-19 mortality in some of the studies. Even though we were able to control for some of the main predictors of COVID-19 mortality (e.g., age, hospital-acquired infections, LTCH residents), we cannot rule out residual confounding. In addition, we considered all patients with a lab-confirmed SARS-CoV-2 diagnosis although we could not confirm that it was the principal reason for the hospitalization. Second, the healthcare administrative and surveillance databases used for this study do not provide detailed information on treatments received by patients. This limitation hampered our ability to examine how evolving standards of care and specific treatments impacted mortality outcomes. Third, we defined our mortality outcome as patients that died within 28 days after admission which may slightly underestimate mortality risk. However, this definition captures close to 90% of the total in-hospital deaths. Additionally, it has the merit of measuring the immediate impact of COVID-19 on deaths more accurately^58^. Finally, the CCM+ data from Ontario did not allow the addition of facility-level variables and vaccine status. We addressed this by using public health unit-level variables to (partially) control for inter-hospital variations. Further, the lack of vaccine status should not affect the results based on the tiny number of fully vaccinated patients (<0.01%) and the similar timeline of vaccination program implemented during the study period.

Strengths of this study includes its representativeness: all hospitalizations in these two provinces are included. This study also adds considerably to the timeline —spanning over three epidemic waves— of COVID-19 inpatient mortality risks and lengths of ICU stay. We controlled for some of the key confounders and results were relatively consistent across provinces operating under different health jurisdiction.

In conclusion, this study demonstrates temporal variability in mortality risk among hospitalized patients that could not be explained by changes in COVID-19 patients’ demographic profiles across epidemic waves. Findings highlight the importance of strategies to buffer against surges in hospital capacity and limiting nosocomial outbreaks to reduce in-hospital mortality risk. As the epidemic continues, there remains a potential for future surges from emergence of new variants, especially if associated with increased virulence, and the potential for waning protection against severity from vaccines; but also the potential for reduction in hospitalization with the scale-up of outpatient therapeutics. Hence, continued monitoring of the evolution of patient outcomes and re-evaluation of the length of ICU stay will be essential to adapt, and inform hospital capacity planning to improve patient outcomes.

## Data Availability

All data used in this study are not available to the public.

## Authors’ contribution

YX, SM, DB, and MMG conceived and designed the study. YX conducted the statistical analysis, conducted the literature search drafted the manuscript. HM supported data curation and cleaning for Ontario. HM, DB, MB, BS, AC, AV, IG, NK, SM, and MMG interpreted results, drafted and edited the manuscript, and critically reviewed it for intellectual content. All authors approved the final version of the manuscript.

## Acknowledgements

This work was supported by McGill’s *Interdisciplinary Initiative for Infection and Immunity (Mi4)* (to MM-G) and a grant from the *Canadian Institutes of Health Research* (to SM). YX is supported by a doctoral award from the *Fonds de recherche du Québec – Santé* (FRQS). BS research program is funded by a *Canada Research Chair (Tier 2) in Economics of Infectious Diseases;* MM-G research program is funded by a *Canada Research Chair (Tier 2) in Population Health Modeling;* NK is supported by a career award from the *Fonds de Recherche Québec – Santé (FRQ-S; Junior 1);* and SM research program is funded by a *Canada Research Chair (Tier 2) in Mathematical Modeling and Program Science*.

