## Supplementary Figures for "Mortality trends and lengths of stay among hospitalized COVID-19 patients in Ontario and Québec (Canada): a population-based cohort study of the first three epidemic waves"

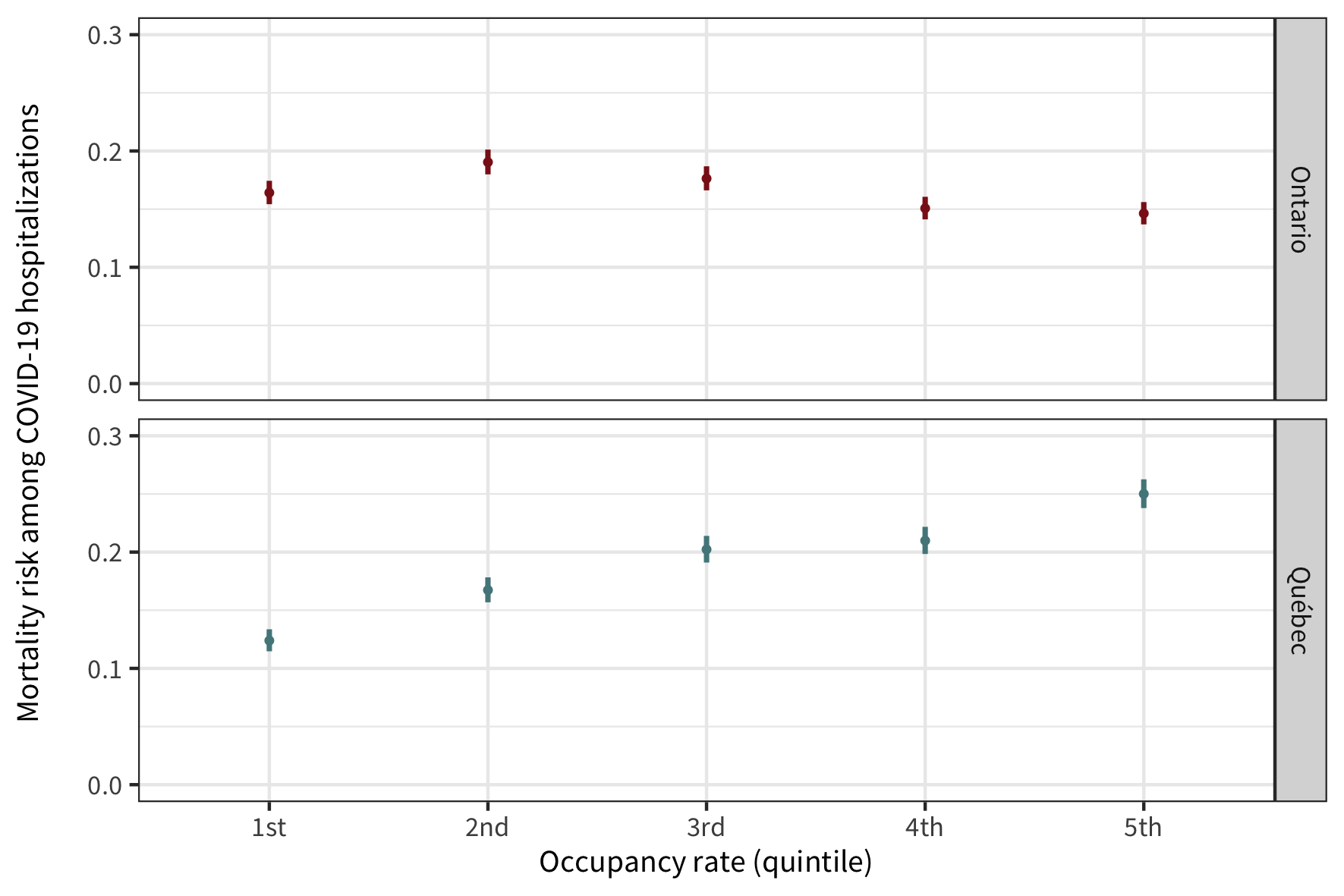
**Figure S1**. Unadjusted mortality risk among patients hospitalized with COVID-19 by quintiles of occupancy with 95% Clopper-Pearson confidence intervals in Ontario (top panel) and Québec (bottom panel) from March 1^st^, 2020 to May 31^st^, 2021. Occupancy rate is ranked separately from the lowest to the highest quintile by public-health unit in Ontario and by facility in Québec.

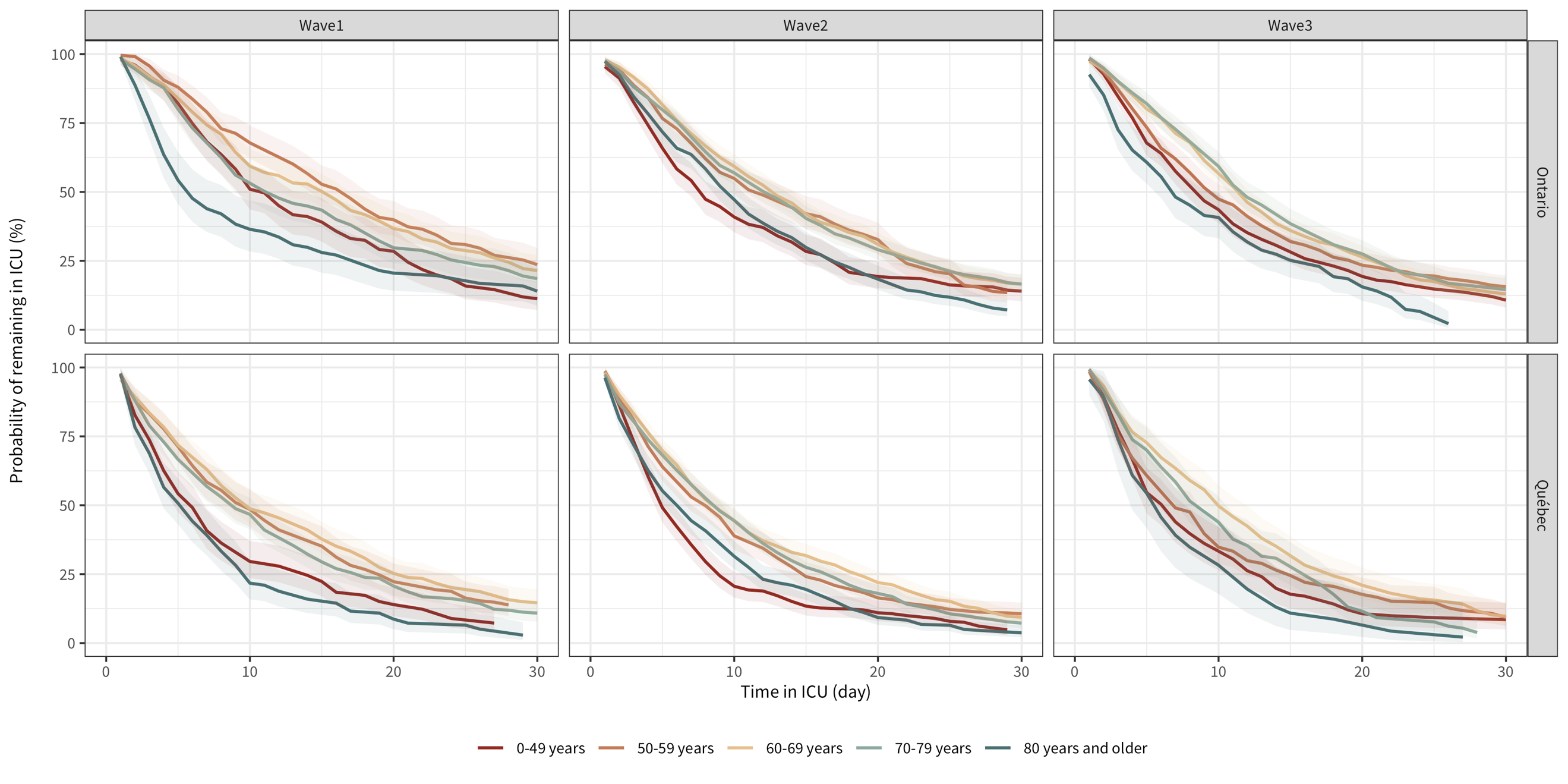
**Figure S2** Kaplan-Meier curves for length of stay in intensive care units (ICU) among COVID-19 ICU inpatients during the 3 waves, stratified by different age groups, in Ontario (top panels) and Québec (bottom panels). Wave1: before June 30^th^, 2020; Wave 2: Aug 23^rd^, 2020 to March 20^th^, 2021; Wave 3: March 21^st^, 2021 to May 31^st^, 2021. Summer lull (July 1^st^, 2020 to Aug 22^nd^, 2020) is excluded due to small number of hospitalizations.

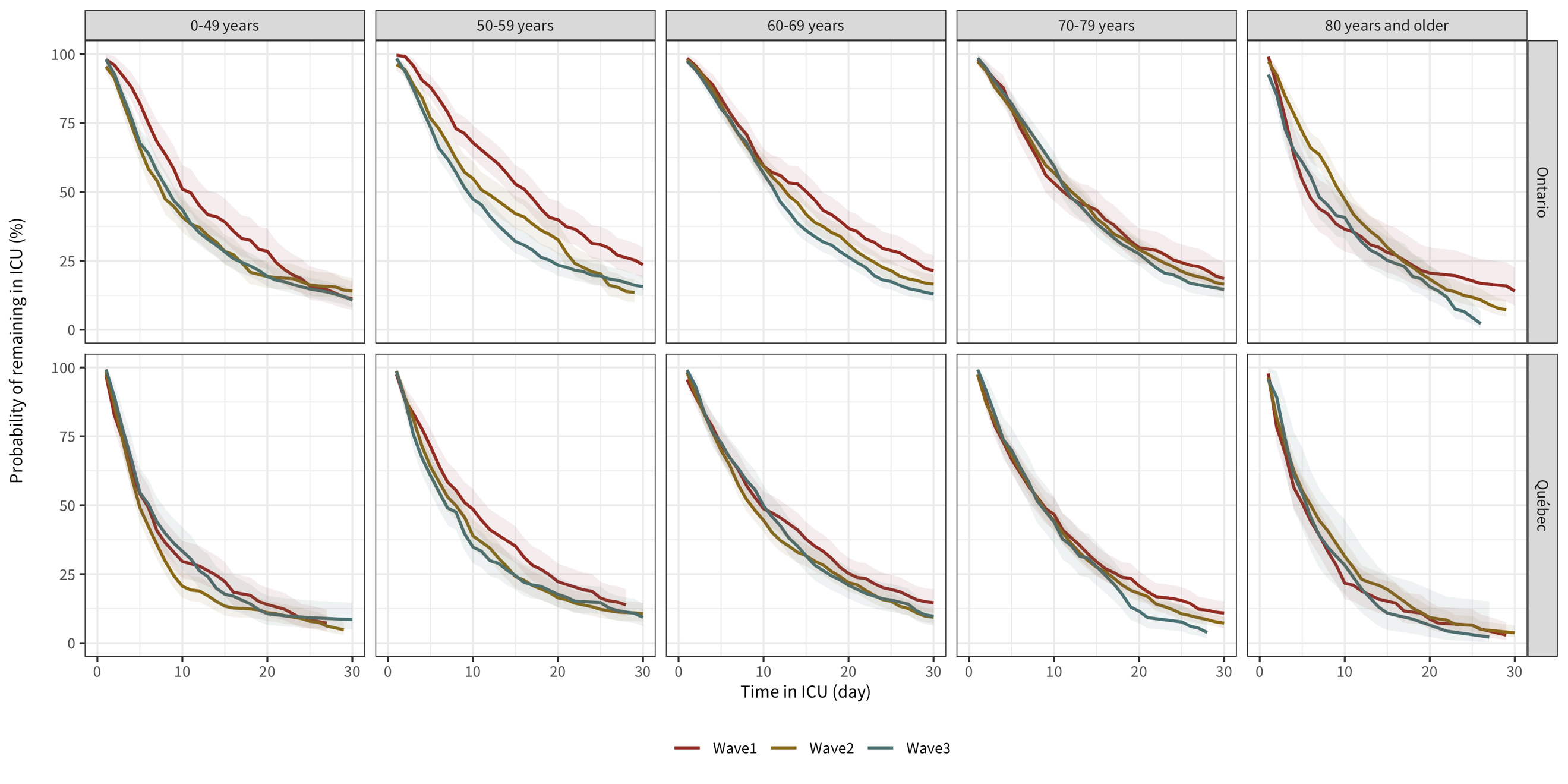
**Figure S3** Kaplan-Meier curves for length of stay in intensive care units (ICU) among COVID-19 ICU inpatients in different age groups, stratified by wave, in Ontario (top panels) and Québec (bottom panels). Wave1: before June 30^th^, 2020; Wave 2: Aug 23^rd^, 2020 to March 20^th^, 2021; Wave 3: March 21^st^, 2021 to May 31^st^, 2021. Summer lull (July 1^st^, 2020 to Aug 22^nd^, 2020) is excluded due to small number of hospitalizations.

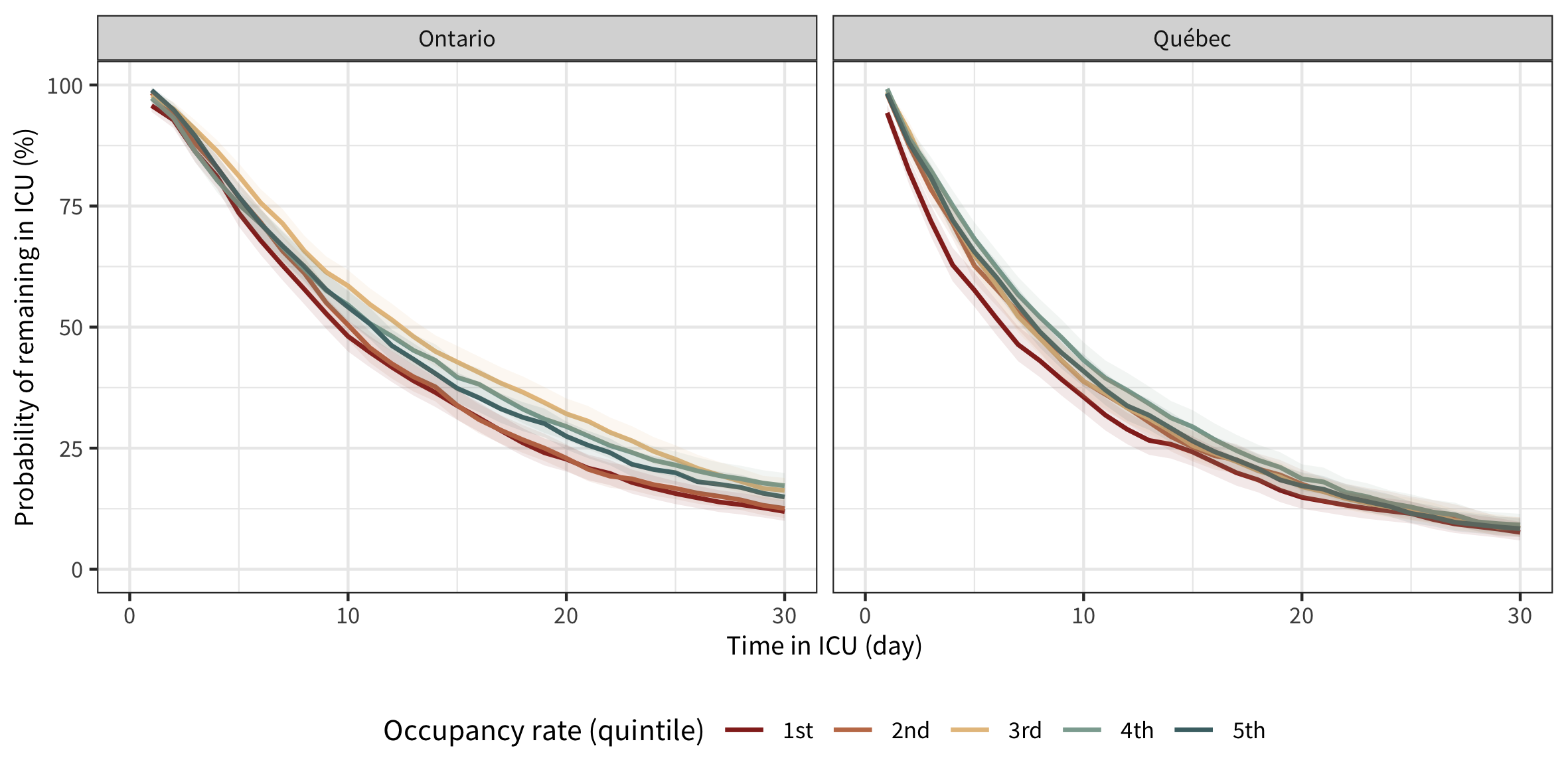
**Figure S4** Kaplan-Meier curves for length of stay in intensive care units (ICU) among COVID-19 ICU inpatients admitted to hospitals in different ranks of ICU occupancy rate, in Ontario and Québec. Wave1: before June 30^th^, 2020; Wave 2: Aug 23^rd^, 2020 to March 20^th^, 2021; Wave 3: March 21^st^, 2021 to May 31^st^, 2021. Summer lull (July 1^st^, 2020 to Aug 22^nd^, 2020) is excluded due to small number of hospitalizations.

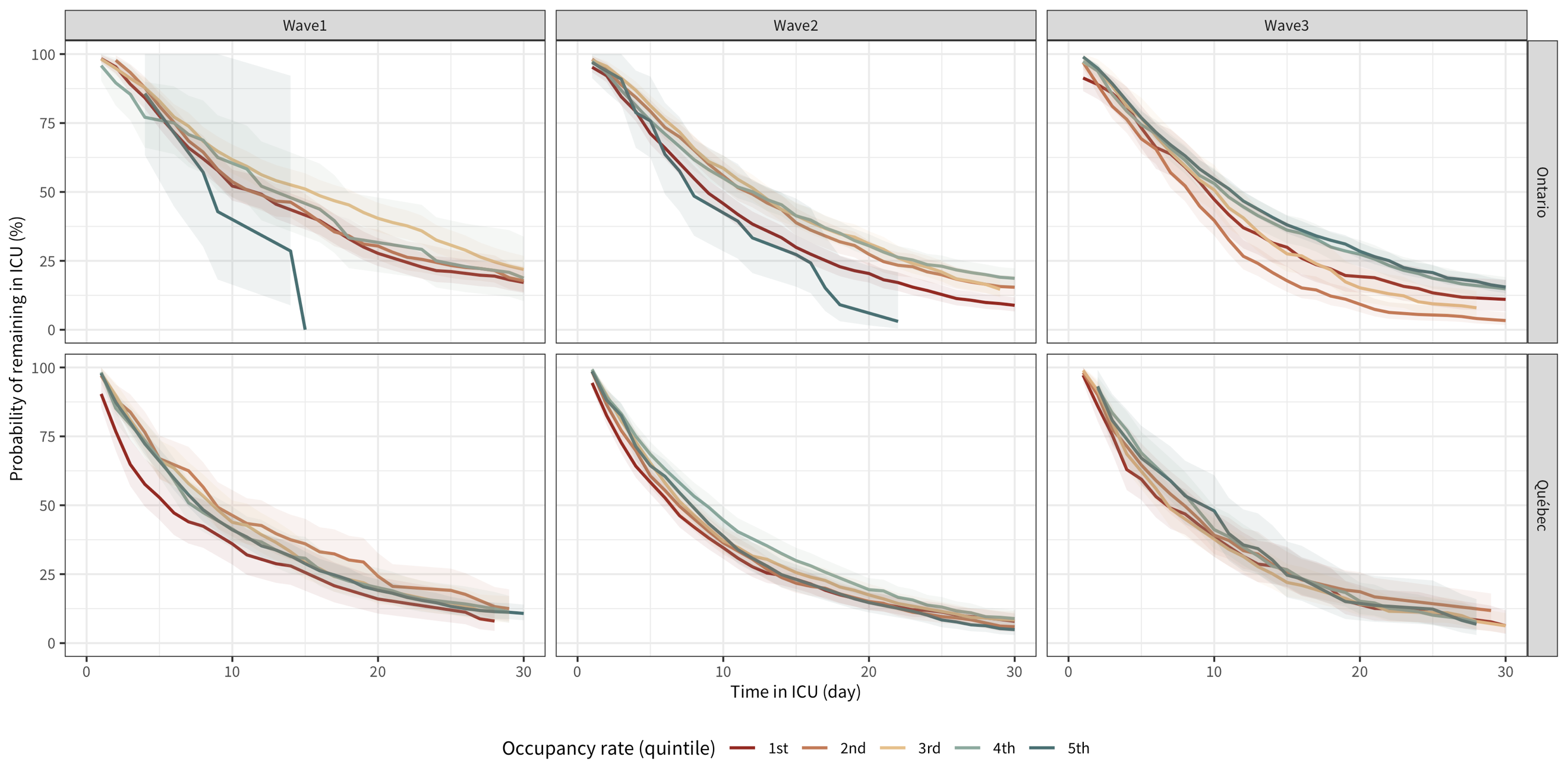
**Figure S5** Kaplan-Meier curves for length of stay in intensive care units (ICU) among COVID-19 ICU inpatients admitted to hospitals in different ranks of ICU occupancy rate, stratified by wave, in Ontario and Québec. Wave1: before June 30^th^, 2020; Wave 2: Aug 23^rd^, 2020 to March 20^th^, 2021; Wave 3: March 21^st^, 2021 to May 31^st^, 2021. Summer lull (July 1^st^, 2020 to Aug 22^nd^, 2020) is excluded due to small number of hospitalizations.

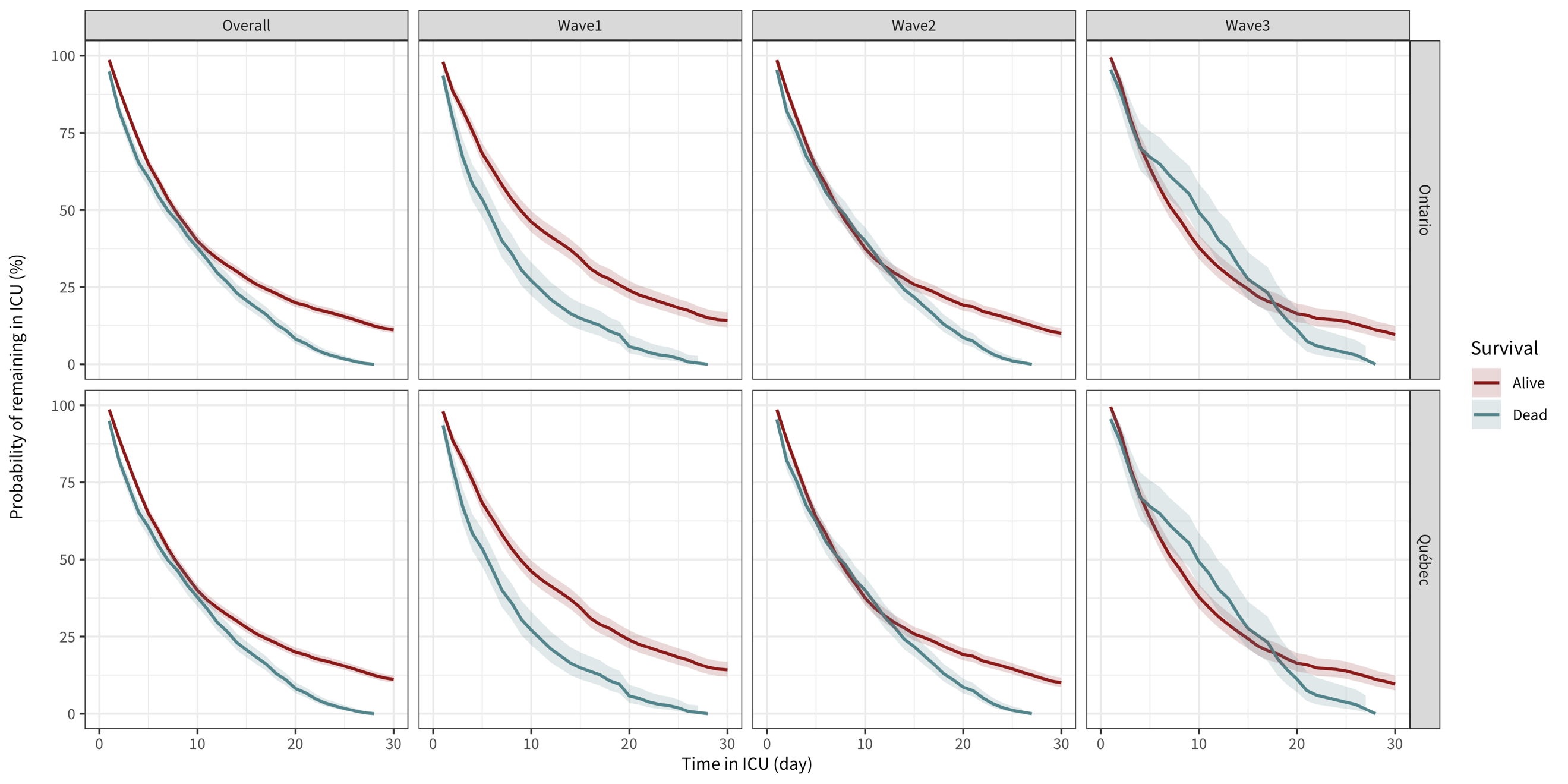

**Figure S6** Kaplan-Meier curves for length of stay in intensive care units (ICU) among COVID-19 ICU inpatients admitted to hospitals stratified by outcome and wave, in Ontario and Québec. Wave1: before June 30^th^, 2020; Wave 2: Aug 23^rd^, 2020 to March 20^th^, 2021; Wave 3: March 21^st^, 2021 to May 31^st^, 2021. Summer lull (July 1^st^, 2020 to Aug 22^nd^, 2020) is excluded due to small number of hospitalizations.

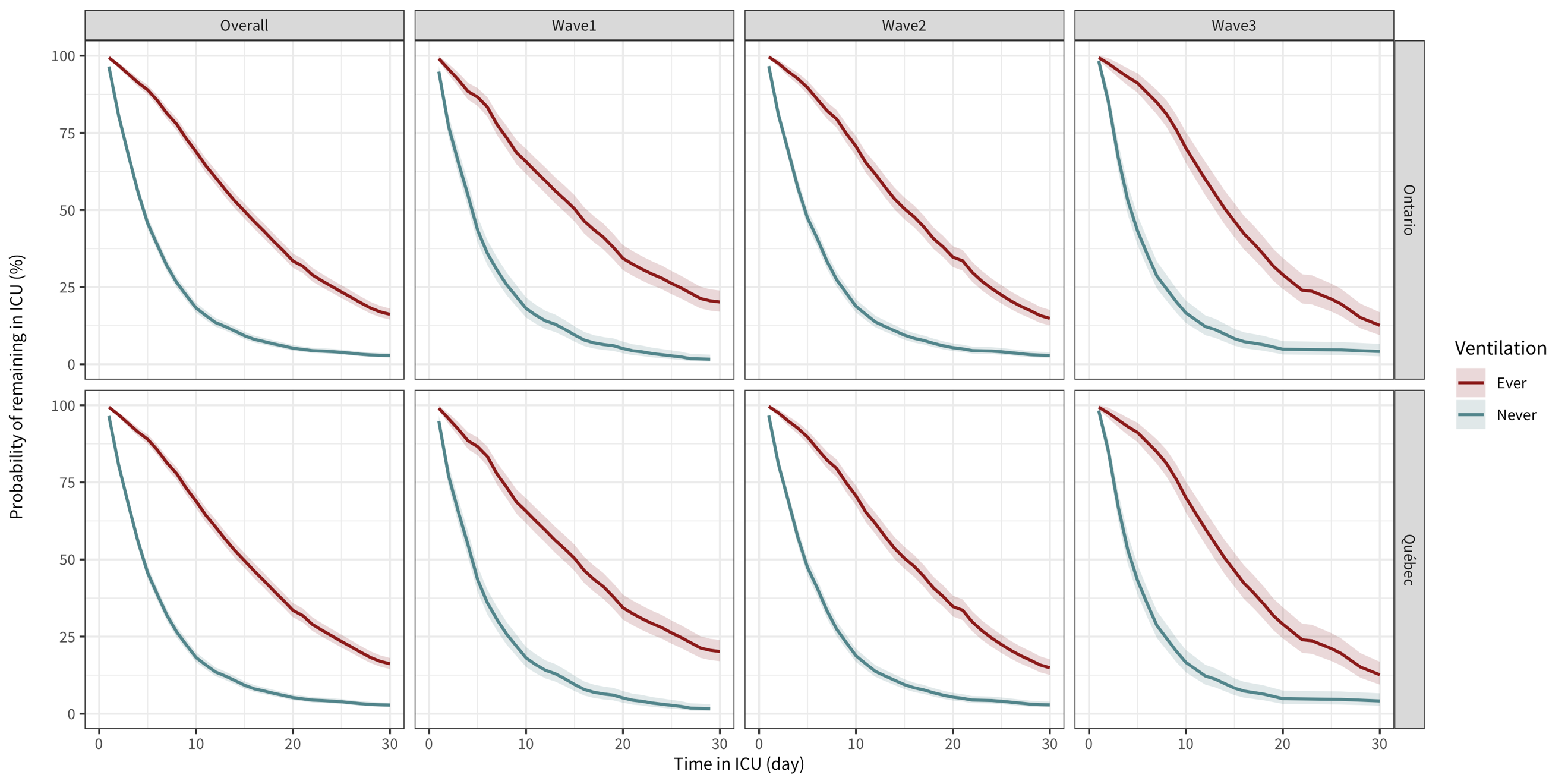

**Figure S7** Kaplan-Meier curves for length of stay in intensive care units (ICU) among COVID-19 ICU inpatients admitted to hospitals stratified by ventilation status and wave, in Ontario and Québec. Wave1: before June 30^th^, 2020; Wave 2: Aug 23^rd^, 2020 to March 20^th^, 2021; Wave 3: March 21^st^, 2021 to May 31^st^, 2021. Summer lull (July 1^st^, 2020 to Aug 22^nd^, 2020) is excluded due to small number of hospitalizations.
