## Supplementary Tables for "Mortality trends and lengths of stay among hospitalized COVID-19 patients in Ontario and Québec (Canada): a population-based cohort study of the first three epidemic waves"

**Table S1** Odds ratios and 95% confidence intervals (CI) for covariates included the logistic regression models^1^ of in-hospital mortality in Ontario and Québec.

|  | Ontario | | | Québec | | |
| --- | --- | --- | --- | --- | --- | --- |
| Covariate | **Odds ratio** | | **95% CI** | **Odds ratio** | | **95% CI** |
| Gender^2^ | |  | | |  | |
| Female | Ref | |  | Not applicable | | |
| Male | 1.4 | | 1.3-1.5 |  |  |  |
| Other | 1.7 | | 0.9-3.0 |  |  |  |
| Living in long-term care facilities | 2.4 | | 2.1-2.7 | 1.7 | | 1.5-2.0 |
| Presumed hospital-acquired infection | 1.5 | | 1.4-1.7 | 1.0 | | 1.0-1.2 |
| Direct admission to ICU | 3.7 | | 3.3-4.1 | 2.5 | | 2.1-3.1 |
| Infected with variants of concern | 2.0 | | 1.7-2.3 | 1.3 | | 1.0-1.7 |
| Occupancy rate | |  | | |  | |
| 1^st^ quintile | Ref | |  | Ref | |  |
| 2^nd^ quintile | 1.1 | | 1.0-1.3 | 1.4 | | 1.2-1.6 |
| 3^rd^ quintile | 1.0 | | 0.9-1.2 | 1.5 | | 1.3-1.7 |
| 4^th^ quintile | 1.0 | | 0.9-1.2 | 1.5 | | 1.3-1.7 |
| 5^th^ quintile | 1.2 | | 1.0-1.4 | 1.6 | | 1.3-1.9 |
| Fully vaccinated before infection^3^ | Not applicable | | | 0.0 | | 0.0-Inf |

^1^ The models were adjusted for quintile of occupancy at time of admission, age (cubic spline with 3 knots at 50, 70, and 80 years), gender (only for Ontario), whether the patient was from long-term care facility, had an hospital-acquired infection, direct admission to the intensive care unit, infection with a variant of concern, full vaccination status (only for Québec), and either facility-level fixed effects (in Québec) or PHU-level fixed effects (in Ontario).

^2^ Due to data limitation, gender is used as an proxy of the biological sex.

^3^ Only 0.1% of the hospitalizations were fully vaccinated before infection

**Table S2** Mean and standard deviation (SD) of length of stay in intensive care units (ICU) by wave, age group, and ICU occupancy rate (quintile ranking) in Ontario and Québec.

|  | Overall  (March 1^st^, 2020 -  May 31^st^, 2021) | | Wave1  (March 1^st^, 2020 -  July 31^st^, 2020) | | Wave2  (August 1^st^, 2020 -  March 20^th^, 2021) | | Wave3  (March 21^st^, 2021 -  May 31^st^, 2021) | |
| --- | --- | --- | --- | --- | --- | --- | --- | --- |
|  | Mean | SD | Mean | SD | Mean | SD | Mean | SD |
| Ontario | | | | | | | | |
| Overall | 17.2 | 18.5 | 19.4 | 19.0 | 17.6 | 20.1 | 15.6 | 15.5 |
| *Occua^b^ 1^st^* | 15.6 | 18.5 | 18.1 | 20.6 | 14.7 | 18.4 | 13.6 | 12.3 |
| *Occup 2^nd^* | 16.0 | 17.2 | 17.9 | 16.3 | 18.5 | 20.9 | 10.5 | 9.0 |
| *Occup 3^rd^* | 19.1 | 19.6 | 22.5 | 22.3 | 18.5 | 19.0 | 13.2 | 11.2 |
| *Occup 4^th^* | 18.1 | 20.0 | 17.4 | 13.9 | 19.2 | 22.1 | 16.7 | 17.0 |
| *Occup 5^th^* | 17.1 | 16.8 | 10.1 | 4.5 | 10.7 | 7.2 | 17.4 | 17.1 |
| Age group | | | | | | | | |
| *0-49 years* | 15.1 | 18.1 | 18.2 | 23.2 | 15.3 | 19.6 | 13.8 | 14.1 |
| *Occup^b^ 1^st^* | 14.1 | 25.2 | 16.0 | 29.4 | 14.1 | 25.6 | 9.7 | 10.2 |
| *Occup 2^nd^* | 12.4 | 13.9 | 14.9 | 13.9 | 15.4 | 18.5 | 8.8 | 8.1 |
| *Occup 3^rd^* | 19.7 | 19.5 | 24.3 | 21.9 | 16.9 | 18.4 | 18.6 | 16.9 |
| *Occup 4^th^* | 14.6 | 16.2 | 5.5 | 4.9 | 14.5 | 17.1 | 15.0 | 15.5 |
| *Occup 5^th^* | 15.1 | 14.8 | 6.0 | Only one observation | 12.7 | 12.8 | 15.3 | 14.9 |
| *50-59 years* | 18.3 | 19.4 | 22.3 | 20.1 | 18.9 | 22.9 | 15.6 | 15.7 |
| *Occup^b^ 1^st^* | 17.7 | 21.6 | 20.7 | 16.1 | 15.9 | 28.4 | 14.2 | 11.8 |
| *Occup 2^nd^* | 15.8 | 15.4 | 17.7 | 14.8 | 19.5 | 20.4 | 10.4 | 7.9 |
| *Occup 3^rd^* | 21.8 | 21.8 | 28.2 | 26.2 | 19.4 | 18.5 | 11.9 | 8.4 |
| *Occup 4^th^* | 19.7 | 20.6 | 18.7 | 14.8 | 21.4 | 23.0 | 18.2 | 18.8 |
| *Occup 5^th^* | 16.9 | 16.9 | 14.5 | 0.7 | 13.0 | 3.8 | 17.0 | 17.2 |
| *60-69 years* | 18.4 | 18.0 | 20.9 | 19.0 | 18.6 | 18.5 | 16.9 | 16.7 |
| *Occup^b^ 1^st^* | 16.9 | 15.2 | 21.6 | 17.7 | 15.4 | 14.4 | 14.3 | 11.8 |
| *Occup 2^nd^* | 17.4 | 18.3 | 21.2 | 19.2 | 18.8 | 20.3 | 11.3 | 11.3 |
| *Occup 3^rd^* | 19.7 | 19.1 | 21.5 | 21.1 | 20.1 | 19.4 | 13.9 | 10.1 |
| *Occup 4^th^* | 19.5 | 19.2 | 16.1 | 11.7 | 20.8 | 19.9 | 18.1 | 19.3 |
| *Occup 5^th^* | 18.4 | 17.7 | 9.3 | 5.5 | 9.9 | 6.7 | 18.8 | 17.9 |
| *70-79 years* | 18.3 | 19.3 | 18.1 | 18.2 | 18.8 | 21.4 | 17.6 | 16.6 |
| *Occup^b^ 1^st^* | 15.7 | 16.4 | 16.6 | 21.5 | 15.2 | 13.9 | 16.5 | 15.1 |
| *Occup 2^nd^* | 18.2 | 16.8 | 19.6 | 16.9 | 20.4 | 19.5 | 12.9 | 8.4 |
| *Occup 3^rd^* | 17.9 | 17.8 | 17.8 | 16.1 | 19.6 | 19.9 | 9.4 | 5.7 |
| *Occup 4^th^* | 20.2 | 24.8 | 21.8 | 19.1 | 21.1 | 28.6 | 18.1 | 16.3 |
| *Occup 5^th^* | 19.5 | 18.8 | 8.0 | Only one observation | 10.8 | 5.6 | 20.0 | 19.1 |
| *80 years and older* | 13.4 | 16.2 | 14.1 | 18.1 | 14.6 | 17.9 | 10.2 | 8.3 |
| *Occup^b^ 1^st^* | 11.7 | 13.9 | 12.0 | 14.9 | 11.6 | 13.6 | 10.2 | 11.6 |
| *Occup 2^nd^* | 14.7 | 20.9 | 11.3 | 12.3 | 17.6 | 25.2 | 8.8 | 7.0 |
| *Occup 3^rd^* | 15.3 | 19.6 | 18.7 | 25.4 | 15.0 | 17.7 | 7.3 | 5.3 |
| *Occup 4^th^* | 13.3 | 11.3 | 17.0 | 13.3 | 14.5 | 12.4 | 9.7 | 7.2 |
| *Occup 5^th^* | 11.1 | 9.2 | No observation | | 4.3 | 1.5 | 11.4 | 9.2 |
| Survival status |  |  |  | |  |  |  |  |
| *Alive* | 19.6 | 21.2 | 23.5 | 22.3 | 20.5 | 23.7 | 16.8 | 17.4 |
| *Dead* | 11.7 | 7.5 | 10.3 | 6.7 | 12.0 | 7.7 | 12.1 | 7.5 |
| Ventilation status |  |  |  | |  |  |  |  |
| *Ever* | 24.1 | 22.9 | 29.4 | 21.4 | 24.0 | 24.6 | 14.3 | 12.2 |
| *Never* | 15.8 | 17.1 | 16.8 | 18.5 | 15.5 | 17.9 | 15.7 | 15.7 |

^a^ Occup = ICU occupancy rate (quintle ranking from lowest to highest)

**Table S2** (cont.)

|  | | Overall  (March 1^st^, 2020 -  May 31^st^, 2021) | | | | Wave1  (March 1^st^, 2020 -  July 31^st^, 2020) | | Wave2  (August 1^st^, 2020 -  March 20^th^, 2021) | | | | Wave3  (March 21^st^, 2021 -  May 31^st^, 2021) | | | |
| --- | --- | --- | --- | --- | --- | --- | --- | --- | --- | --- | --- | --- | --- | --- | --- |
|  | | Mean | | SD | | Mean | SD | Mean | | SD | | Mean | | SD | |
| Québec | | | | | | | | | | | | | | | |
| Overall | | 12.9 | | 15.2 | | 13.5 | 15.2 | 12.6 | | 15.5 | | 12.6 | | 14.1 | |
| *Occup^a^ 1^st^* | | 11.9 | | 15.9 | | 12.5 | 19.2 | 11.8 | | 15.4 | | 11.9 | | 14.1 | |
| *Occup 2^nd^* | | 13.2 | | 16.4 | | 15.6 | 16.3 | 11.9 | | 15.3 | | 14.8 | | 19.1 | |
| *Occup 3^rd^* | | 12.9 | | 15.4 | | 14.0 | 15.5 | 13.3 | | 17.1 | | 11.3 | | 10.6 | |
| *Occup 4^th^* | | 13.8 | | 15.4 | | 14.0 | 17.3 | 14.0 | | 15.7 | | 12.7 | | 12.4 | |
| *Occup 5^th^* | | 12.5 | | 12.7 | | 12.8 | 12.6 | 11.9 | | 13.1 | | 12.6 | | 12.0 | |
| Age group | | | | | | | | | | | | | | | |
| *0-49 years* | | 10.3 | | 13.9 | | 10.9 | 15.3 | 9.4 | | 12.1 | | 11.6 | | 15.2 | |
| *Occup^b^ 1^st^* | | 8.5 | | 17.2 | | 11.7 | 29.0 | 6.8 | | 12.2 | | 9.7 | | 13.8 | |
| *Occup 2^nd^* | | 11.0 | | 15.7 | | 9.1 | 6.7 | 9.5 | | 13.5 | | 15.5 | | 23.0 | |
| *Occup 3^rd^* | | 9.4 | | 9.2 | | 9.2 | 8.6 | 9.3 | | 9.6 | | 9.9 | | 9.4 | |
| *Occup 4^th^* | | 9.8 | | 11.0 | | 7.9 | 7.6 | 11.4 | | 13.0 | | 8.4 | | 8.9 | |
| *Occup 5^th^* | | 14.0 | | 13.5 | | 13.8 | 13.3 | 12.1 | | 10.6 | | 19.6 | | 19.4 | |
| *50-59 years* | | 14.3 | | 18.8 | | 15.3 | 16.7 | 14.9 | | 22.4 | | 12.5 | | 14.2 | |
| *Occup^b^ 1^st^* | | 14.9 | | 21.2 | | 17.7 | 20.1 | 15.3 | | 24.7 | | 13.0 | | 15.9 | |
| *Occup 2^nd^* | | 12.8 | | 20.4 | | 13.6 | 11.2 | 11.7 | | 24.4 | | 14.3 | | 17.9 | |
| *Occup 3^rd^* | | 14.9 | | 19.4 | | 19.4 | 24.1 | 16.2 | | 21.7 | | 10.5 | | 10.9 | |
| *Occup 4^th^* | | 14.1 | | 16.3 | | 11.8 | 12.9 | 15.0 | | 18.1 | | 13.8 | | 14.4 | |
| *Occup 5^th^* | | 15.0 | | 16.9 | | 14.8 | 14.6 | 18.2 | | 24.1 | | 10.8 | | 10.3 | |
| *60-69 years* | | 14.8 | | 15.2 | | 15.8 | 15.1 | 14.1 | | 15.0 | | 15.1 | | 16.2 | |
| *Occup^b^ 1^st^* | | 13.9 | | 14.2 | | 18.0 | 15.1 | 13.2 | | 13.5 | | 13.8 | | 16.2 | |
| *Occup 2^nd^* | | 16.6 | | 18.0 | | 23.7 | 22.9 | 13.6 | | 11.9 | | 18.3 | | 24.1 | |
| *Occup 3^rd^* | | 14.1 | | 13.7 | | 17.1 | 14.1 | 12.6 | | 14.1 | | 14.5 | | 12.2 | |
| *Occup 4^th^* | | 17.0 | | 18.1 | | 15.4 | 14.3 | 18.7 | | 21.4 | | 15.2 | | 13.4 | |
| *Occup 5^th^* | | 12.3 | | 10.8 | | 12.7 | 11.6 | 12.1 | | 10.2 | | 10.3 | | 6.8 | |
| *70-79 years* | | 13.2 | | 14.5 | | 14.0 | 15.8 | 13.1 | | 14.7 | | 11.5 | | 9.4 | |
| *Occup^b^ 1^st^* | | 12.6 | | 13.1 | | 10.1 | 12.2 | 13.5 | | 13.9 | | 12.1 | | 8.9 | |
| *Occup 2^nd^* | | 12.7 | | 13.0 | | 17.2 | 15.1 | 11.9 | | 12.9 | | 12.3 | | 10.9 | |
| *Occup 3^rd^* | | 14.1 | | 18.4 | | 12.3 | 13.9 | 15.4 | | 20.8 | | 10.6 | | 9.8 | |
| *Occup 4^th^* | | 13.8 | | 15.3 | | 17.1 | 23.8 | 12.8 | | 11.1 | | 11.1 | | 6.4 | |
| *Occup 5^th^* | | 12.7 | | 11.7 | | 13.8 | 13.1 | 11.4 | | 9.9 | | 11.3 | | 9.6 | |
| *80 years and older* | | 9.4 | | 10.6 | | 8.7 | 9.5 | 9.9 | | 11.3 | | 8.7 | | 8.5 | |
| *Occup^b^ 1^st^* | | 9.4 | | 11.9 | | 6.6 | 7.1 | 10.0 | | 12.8 | | 7.6 | | 8.5 | |
| *Occup 2^nd^* | | 9.8 | | 11.9 | | 7.4 | 6.6 | 10.8 | | 13.6 | | 7.5 | | 5.2 | |
| *Occup 3^rd^* | | 9.4 | | 7.7 | | 9.2 | 6.4 | 9.9 | | 8.8 | | 7.9 | | 4.9 | |
| *Occup 4^th^* | | 11.7 | | 12.1 | | 17.2 | 18.3 | 10.4 | | 9.1 | | 10.1 | | 12.7 | |
| *Occup 5^th^* | | 7.0 | | 8.2 | | 6.5 | 5.4 | 7.4 | | 10.6 | | 18.0 | | Only one observation | |
| Survival status |  | |  | |  | | | |  | |  | |  | |  |
| *Alive* | 14.0 | | 16.9 | | 15.3 | | 16.7 | | 13.7 | | 17.5 | | 13.0 | | 15.2 |
| *Dead* | 9.3 | | 6.9 | | 8.0 | | 6.5 | | 9.5 | | 6.8 | | 10.9 | | 7.3 |
| Ventilation status |  | |  | |  | | | |  | |  | |  | |  |
| *Ever* | 19.9 | | 18.0 | | 19.9 | | 17.1 | | 20.5 | | 19.8 | | 18.6 | | 14.9 |
| *Never* | 7.8 | | 10.0 | | 7.4 | | 9.8 | | 7.8 | | 9.5 | | 8.0 | | 11.5 |

^a^ Occup = ICU occupancy rate (quintle ranking from lowest to highest)
