## Supplementary Text for "Mortality trends and lengths of stay among hospitalized COVID-19 patients in Ontario and Québec (Canada): a population-based cohort study of the first three epidemic waves"

**Missing dates related to hospitalization**

Date imputation was conducted before any inclusion and exclusion process. Therefore, the proportion missing mentioned in this document is based on the raw dataset (the whole IPHIS dataset transferred on August 15^th^, 2021). According to the nature of administrative dataset, a larger proportion of missing value occurs within more recently entered cases. Individuals marked as “hospitalized” and had at least one date related to hospitalization were regarded as “hospitalizations”.

Figure 1 shows the flow char of the data processing procedure. Of the 28247 records, 62 missing dates of specimen collection were replaced with episode date. Missing dates of hospital admission (523 / 28247, 1.9%) was were with date of specimen collection. For missing dates of hospital discharge (6350 / 28247, 22.5%), they were imputed using date of death (if the patient decreased, 885 / 6350, 13.9%), the latest date among date of ICU discharge, and date of the end of ventilation (if case was resolved, 434 / 6350, 6.9%), or the date of data cut-off of the dataset (if case was not resolved, 103 / 6350, 1.6%). After imputation, 17.4% (4929 / 28247) of discharge date were still missing. Missing dates of ICU admission (38 / 5524, <0.1%) were substituted using date of hospital admission if the patient had an indication of ever in ICU. Missing dates of ICU discharge (1025 / 5524, 18.6%) were filled following the same logic as date of hospital discharge (284 / 1025 (27.7%) using date of death, 137 / 1025 (13.4%) using maximum date, and 27 / 1025 (2.6%) using data cut-off date). After processing, there were 10.4% (577 / 5524) missing date of ICU discharge remaining. All mssing dates of deaths (21 / 5407, <0.1%) were imputed with the latest date among date of hospital discharge, date of ICU discharge, and date of the end of vitalization.

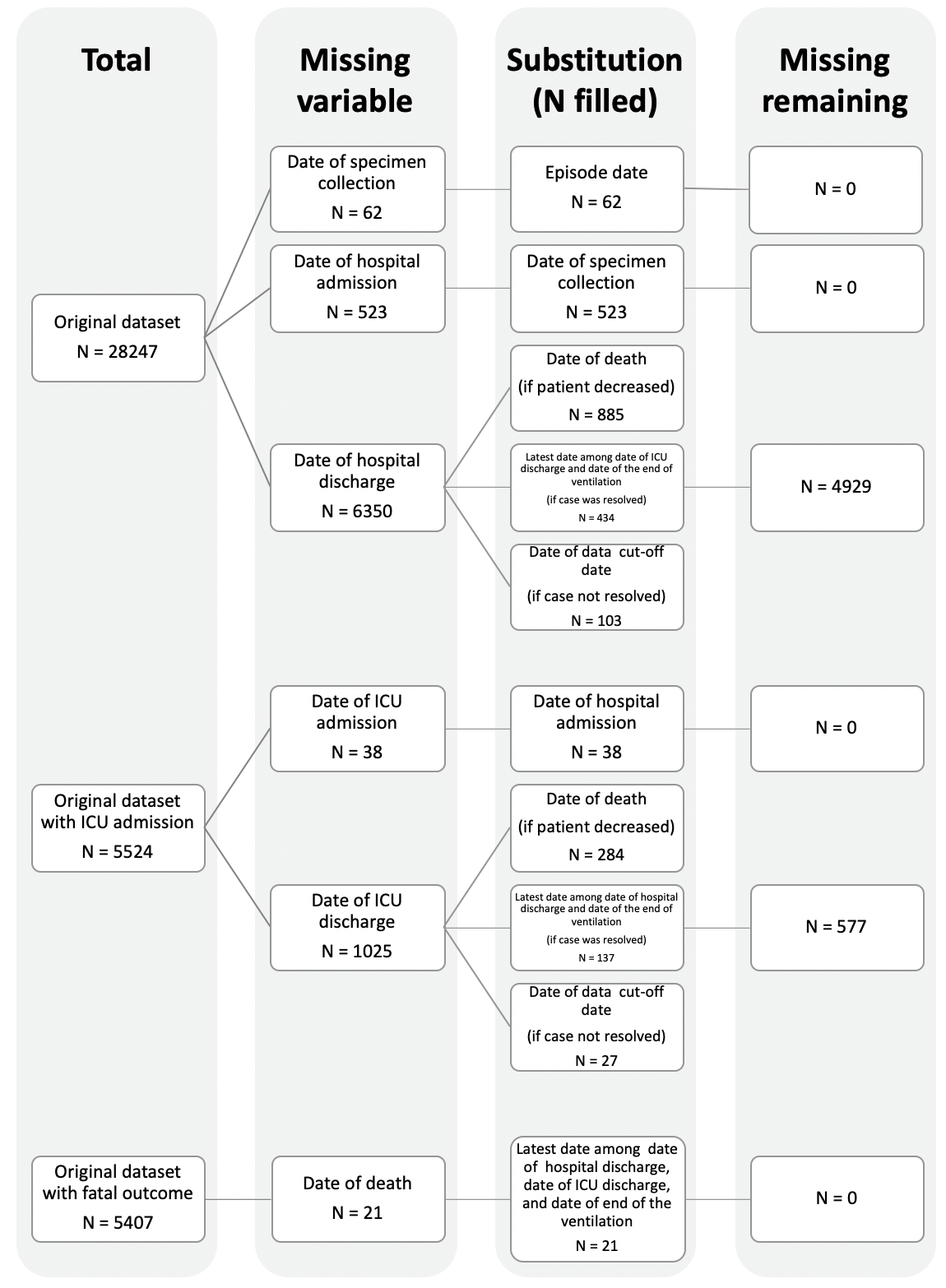
**Figure 1** Flow chart on data processing procedure before analysis in Ontario.

**Missing Public Health Units**

There were 1.5% (434 / 28247) records missing Public Health Units (PHU) of residence, they were assigned the PHU where the patients were diagnosed.

**Missing continuous age**

Starting in July 2021, IPHIS reports only reports the 10-year age group to which the individuals were belonged to and classifies all cases older than 80 years old into one group. According to a study published using Ontario data (1), survival rate decreased with age (presented in 2-year age group). Therefore, we looked into the latest IPHIS report available with a 2-year age group (this is the most precise age variable in IPHIS reports) to match with the August 15 dataset. The IPHIS report extracted on June 28^th^, 2021 was used.

Based on the fact that no specific identification variable can be utilized to link the two datasets, we matched on the 10-year age group, the episode date, the date of specimen collection, the PHU of diagnosis, gender, the dissemination area of residence, the date of admission, the date of death, the date of discharge, the date of create of record, the date of report, and the date of information collection. After matching, 270 (<0.1%) of the records in the August 15 dataset failed to find a match. All of the records were in the “80 plus” group. Therefore, those records were imputed with the mean age of people older than 80 years.

Notably, in our study, age is modeled as a continuous variable. Unfortunately, IPHIS reports do not contain the exact age of the patient. As mentioned above, 2-year age group is the finest age information available. As such, we assigned the continuous age of each patient using the mean of the 2-year age group. For example, a patient in the 30-31 age group was given an age of 30.5. Additionally, those who were older than 100 years old were categorized in to one group as “100 Plus”. To make the model feasible, we imputed the age with 100.5.
